# Perinatal outcomes following periconceptual steroid exposure: a population-wide cohort study

**DOI:** 10.64898/2025.12.22.25342798

**Authors:** Anthea Lindquist, Anna Forsythe, Richard Hiscock, Stephen Tong, Susan Walker, Amber Kennedy, Natasha Pritchard, Elizabeth McCarthy, Hannah Gordon, Jessica Atkinson, Beverley Vollenhoven, Mark Green, Catharyn Stern, Roxanne Hastie

## Abstract

**Importance:** Glucocorticoid steroids are increasingly prescribed during the periconceptual period with the hypothesis that they reduce intrauterine inflammation and improve pregnancy rates. There is no robust evidence to support this practice, and the potential harm has not been well characterised.

**Objective:** To examine the risk of adverse perinatal outcomes associated with non-medically indicated glucocorticoid steroid use during the periconceptual period.

**Design:** Population-wide linked retrospective cohort study.

**Setting:** Victoria, Australia.

**Participants:** After excluding women with medical indications for steroid use (autoimmune disease, chronic asthma and previous organ transplant), our total cohort included 805,353 births between 2009 and 2021.

**Exposure:** Prescriptions of glucocorticoid steroids dispensed during the periconceptual period (12 weeks prior to conception – end of first trimester).

**Main Outcome(s) and Measure(s):** Four primary outcomes were examined – spontaneous preterm birth before 37 completed weeks’ gestation, small for gestational age (<10th birthweight centile), major congenital abnormality and perinatal mortality. A doubly robust inverse probability weighted regression adjustment model was used to estimate the association between glucocorticoid steroid exposure and outcomes and presented as adjusted relative risks (aRR) with corresponding 95% confidence intervals (95% CI).

**Results:** There were 12,301 (1.5%) pregnancies exposed to glucocorticoid steroids during the periconceptual period and 793,052 unexposed. Among the steroid-exposed cohort, major congenital abnormalities occurred in 4.5% of pregnancies, compared with 3.5% among those unexposed to steroids. This resulted in a 23% increased risk of major congenital abnormality (aRR 1.23, 95%CI 1.13-1.34). There were no significant associations between steroid exposure and spontaneous preterm birth (2.4 vs 2.5%; aRR 1.05, 95%CI 0.93-1.19), small for gestational age neonates (9.4 vs 9.3%; aRR 1.02, 95%CI 0.97-1.07) or perinatal mortality (0.5 vs 0.7%; aRR 1.05, 95%CI 0.87-1.26).

**Conclusions and Relevance:** In our cohort, periconceptual steroid exposure was associated with an increased risk of major congenital abnormality. In the absence of clear clinical indications, avoiding the prescription of periconceptual steroids is critically important.

**KEY POINTS:** *Question:* What are the risks of adverse perinatal outcomes among patients prescribed glucocorticoid steroids during the periconceptual period?

*Findings:* Periconceptual steroid use is associated with an increased risk of major congenital abnormality.

*Meaning:* In the absence of an indication, periconceptual steroid use does not have clear evidence of benefit, there is now also evidence of significant harm and this practice should not be recommended.

## INTRODUCTION

In an attempt to improve pregnancy rates, adjunct medications are being increasingly prescribed to women undergoing fertility treatment^1–6^. They aim to mitigate the drivers of subfertility, such as inflammation. For instance, glucocorticoid steroids mimic endogenous steroid hormones and can reduce inflammation by suppressing the body’s immune response ^7–9^. This class of medicines has been adopted into guidelines worldwide for peri-implantation pregnancy support for women undergoing fertility treatment ^10–12^.

Both dexamethasone, a potent and long-acting glucocorticoid and prednisolone, a short-acting glucocorticoid, can reduce inflammation^7^. Early studies suggest that steroids can reduce inflammation in the intrauterine environment, suppress natural killer cell activity and in turn, improve the likelihood of ongoing pregnancy ^13,14^. Yet the evidence supporting the use of steroid medications in the setting of conception is insufficient, conflicting or based on animal models only^15–17^. A 2022 Cochrane systematic review of 16 randomised control trials found no evidence that glucocorticoid treatment during the peri-implantation period improved pregnancy rates or livebirth rates, nor reduced the risk of miscarriage ^9^.

In addition to this lack of benefit, data from observational studies have repeatedly shown an increased risk of congenital abnormalities with peri-conceptual steroid use^18,19^. However, this risk has been consistently attributed to the likely underlying medical conditions (eg. autoimmune conditions) among patients taking steroids.

Using population-wide linked data, our study aimed to examine the risk of adverse perinatal outcomes among patients prescribed glucocorticoids during the periconceptual period. Uniquely, we were able to identify and exclude patients with a known medical indication for steroid use, including autoimmune disease, chronic asthma and previous organ transplant.

## METHODS

### Study population

Our retrospective cohort study included all singleton births without a medical indication for steroid use between 2009 and 2021 in the state of Victoria, Australia. Statewide pregnancy, perinatal and birth data were obtained from the Victorian Perinatal Data Collection (VPDC). Population-wide birth records were linked with data on medication prescription (the Pharmaceutical Benefit Scheme – PBS), hospital admission, emergency department presentations and the National Death Index. Data linkage was performed by the Centre for Victorian Data Linkage and the Australian Institute of Health and Welfare. Deidentified linked data were provided to the researchers.

To overcome the potential for confounding by indication, patients were excluded if they had any documented medical indication for prescription steroid use. Indications for steroid use included any autoimmune disease, severe asthma (requiring hospital admission or presentation to the emergency department for treatment) and previous organ transplantation. These conditions were identified using ICD-10 codes (Supp File 1, Table 1) that were documented during any pregnancy episode, hospital admission or emergency department presentation throughout the study period. Other exclusion criteria included missing gestational age, periconceptual exposure to other adjunct prescription medications (tetracycline antibiotics, enoxaparin and oral/per vaginal progesterone) and exposure to medications or conditions known to have teratogenic effects (Supp File 1, Table 2).

### Outcome, exposure and covariates

Four primary outcomes were examined – spontaneous preterm birth before 37 completed weeks’ gestation, small for gestational age, major congenital abnormalities and perinatal mortality. Spontaneous preterm birth was ascertained from VPDC birth records and based on gestational age at birth and onset of labour, in addition to relevant ICD-10 codes (O60: preterm labour and delivery, P072: extreme immaturity (<28 weeks’ gestation), or P073: other preterm infants (28 to <37 weeks gestational age). Small for gestational age was defined as <10th centile based on Australian Institute of Health and Welfare published birthweight percentiles^20^. Major congenital abnormalities were identified via VPDC and hospital admission data using the EUROCAT definition of abnormality^21^. Perinatal mortality included stillbirth after 20 weeks’ gestation or neonatal death within 6 weeks of birth and was captured via VPDC and the National Death Index.

Maternal exposure to prescription steroid was defined as at least one PBS prescription and pharmacy dispensing record of prednisolone and/or dexamethasone during the periconceptual period (12 weeks prior to conception date through to the end of first trimester).

Maternal and pregnancy characteristics considered as potential confounders included body mass index (BMI), maternal age, socioeconomic position (SEIFA), parity, conception via assisted reproductive techniques, pre-existing diabetes, pre-existing hypertension and previous miscarriage or preterm birth. Post-exposure characteristics including birthweight, gestational age at birth and smoking in pregnancy were accounted for by regression adjustment. These data were captured from ICD-10 codes and designated data points from VPDC and hospital admission data. Socioeconomic position was defined using postcode-derived Socio-Economic Index for Areas, with quintile five classified as least deprived^22^. Conception via assisted reproductive technique was captured from VPDC and hospital admission data.

### Handling of missing data

Relevant covariates were examined for proportion and pattern of missingness, with <10% any-missingness across the covariates of BMI, maternal age, SEIFA, smoking status, parity, birthweight and mode of conception (Table 1). All covariates were missing <1% of data except for BMI (6.4%). There were no missing exposure or outcome data. Under the assumption that missing data were missing at random, all covariates with missing values were imputed using multiple imputation in combination with bootstrapping^23^, as described previously ^24^. In brief, the bootstrap sample was set at 1000, imputing two datasets for each bootstrapped sample. The distribution of the imputed data was compared with non-imputed data to ensure the adequacy of imputation (Supp File 2, Table 3, Figure 1).

### Statistical analyses

A prespecified statistical analysis plan was agreed upon by all authors prior to commencing analysis (Supp File 3). All analyses were performed using StataMP version 18^15^. Demographic and clinical characteristics were described by exposure status – periconceptual steroid exposure vs no periconceptual steroid exposure.

Following imputation and bootstrapping, the association between steroid dispensation and each outcome was assessed using a doubly robust inverse probability weighted regression adjustment model (IPWRA). This approach combines two models – an inverse-probability-weighting model that predicts the likelihood of exposure, and an adjusted regression model for the outcome^25,26^. Covariates for both models were considered *a priori* by our expert multidisciplinary team and via the use of Direct Acyclic Graphs (Supp File 3). The propensity score/selection model for the exposure to steroids included maternal BMI, maternal age, SEIFA quintile and pre-existing diabetes for all models. Positivity and covariate balance in the propensity score models was assessed by exposure status across 10% of all datasets, with a standardized mean difference of <0.1 considered adequate (Supp File 2, Table 3, Figure 2).

All outcome models included maternal BMI, maternal age, SEIFA quintile, smoking and parity. Additional covariates were included in the outcome models for preterm birth (previous preterm birth), fetal growth restriction (previous fetal growth restriction and pre-existing hypertension) and perinatal mortality (current pregnancy birthweight and pre-existing hypertension). The average treatment effect across the bootstrapped datasets was calculated using the BootImpute von Hippel method using a one-way ANOVA model to determine within- and between-dataset variance, standard errors, and associated 95% confidence intervals. Clustered standard errors by maternal identifier were used for all models to account for non-independence of outcomes among babies of the same mother.

### Sensitivity and sub-group analyses

Comparative analyses were conducted on complete cases (no missing data) and for Inverse Probability Weighting and Regression Adjustment models separately.

Sensitivity analyses were performed examining each type of steroid (dexamethasone only (N=12,006) and prednisolone only (N=268)). We also performed sub-group analyses restricting our cohort to only pregnancies conceived via assisted reproductive technique (N=26,576).

Ethical approval was granted by the Mercy Health Ethics Committee [HREC 2020-048] on 22^nd^ July 2022. Due to the deidentified nature of the data, patient consent was not required. This research adheres to the Strengthening the Reporting of Observational Studies in Epidemiology (STROBE) guidelines.

## RESULTS

After excluding women with a medical indication for steroid use (N=29,774) and applying other exclusion criteria (N=45,058), our cohort included 805,353 births. There were 12,301 pregnancies (1.5%) exposed to steroid medication during the periconceptual period and 793,052 unexposed (Figure 1). The average (mean) number of prescriptions per pregnancy in the overall exposed group was 1.2 (range 1-16). Among our exposed cohort, 12,033 (97.8%) were exposed to prednisolone and 295 (2.4%) to dexamethasone (27 (0.2%) were exposed to both medications). The average number of scripts per exposed pregnancy was 1.2 for prednisolone (range 1-16) and 1.4 for dexamethasone (range 1-8). The median dose was 25mg/day (range 0.5-1000mg) for prednisolone exposed pregnancies (71.7% of cohort) and 4.0 mg/day (range 0.5-40 mg) for dexamethasone (51.4% of cohort).

Comparing women who dispensed a steroid prescription during the periconceptual period with those that did not, women in the steroid group were more likely to be Australian-born, of higher socioeconomic position (5^th^ quintile), current/past smokers, multiparous, conceived via assisted reproductive techniques and have reported previous pregnancy complications such as recurrent miscarriage, preterm birth and small-for-gestational age infant (<10^th^ centile) (Table 1). Hypertensive disorders of pregnancy were also more common in the index pregnancy among those exposed to steroids.

Median gestational age at birth was 38 completed weeks in both exposure groups (38^+6^ unexposed and 38+^4^ exposed), but early term birth (37-38 weeks) was more common in the steroid exposed cohort (37.1 vs 29.6%). The exposure cohorts had comparable mean birthweights (3373g (SD 556) in unexposed vs 3325 (SD 580) exposed) (Table 1).

Congenital abnormalities (any) occurred in 7.6% of steroid-exposed pregnancies, compared with 6.3% of those unexposed. The rate of major abnormalities was 4.5% for steroid-exposed and 3.5% for steroid unexposed pregnancies (Table 2). This resulted in both a significantly increased crude risk (RR 1.29; 95%CI 1.18-1.40) and adjusted risk (RR 1.23; 95%CI 1.13-1.34) of major congenital abnormalities. Major congenital abnormalities were dominated by circulatory (28.3%), musculoskeletal (24.8%) and urogenital (14.3%) malformations. Chromosomal abnormalities accounted for 3.6% (1,027) of major abnormalities.

Investigating other adverse perinatal outcomes, we found no evidence of association between periconceptual steroid use and spontaneous preterm birth (2.5 vs 2.4%; aRR 1.05, 95%CI 0.93-1.19), small for gestational age (9.4 vs 9.3%; aRR 1.02, 95%CI 0.97-1.07) or perinatal mortality (0.5 vs 0.7%; aRR 1.05, 95%CI 0.87-1.26) (Table 2, Figure 2).

Investigating type of steroid prescribed, we found that prednisolone was associated with a 22% increased risk of major congenital abnormality (aRR 1.23, 95%CI 1.13-1.34) (Supp File 4, Table 4). We found no evidence for an association between dexamethasone alone and risk of major congenital abnormalities (aRR 1.21, 95%CI 0.58-2.52), although exposure numbers were small.

Next, we restricted our cohort to pregnancies conceived following ART (n=26,576) (Supp File 4, Table 5). Among this cohort, steroid-exposure was associated with a 55% higher risk of major congenital abnormality (aRR 1.55; 95%CI 1.10-2.20) compared with unexposed pregnancies. The risks of preterm birth (aRR 0.81; 95%CI 0.44-1.49), small for gestational age (aRR 0.94; 95%CI 0.71-1.24) and perinatal mortality (aRR 1.49; 95%CI 0.82-2.73) were not significantly different among this cohort between women prescribed steroids and those not.

Complete case analysis, IPW model and regression adjusted models revealed similar results to our primary IPWRA model (Supp File 4, Table 6).

## DISCUSSION

Utilising a large statewide data linkage, we have demonstrated that periconceptual steroid use is associated with a significantly increased relative risk of major congenital abnormalities, even after the exclusion of patients with a clear medical indication for use. Our findings suggest the risk of major abnormalities is even more pronounced (55% higher) among pregnancies conceived with assisted reproductive techniques. While peri-conceptual steroids continue to be offered to patients under the auspices of pregnancy support, our findings may have substantial implications for clinical practice and patients worldwide.

Outside of proven clinical indications, steroid use in early pregnancy is limited to the setting of ART and fertility treatment. There are myriad fertility clinic websites and patient information resources online that provide a cautious, but not discouraging, recommendation for glucocorticoid use during early pregnancy during and after fertility treatments. The premise offered is to reduce pregnancy loss and promote ongoing pregnancy^27^. However, the evidence of benefit is limited.

A previous matched case-control study examined the benefit of combined co-treatment with aspirin, doxycycline and prednisolone, with or without oestradiol patches on livebirth rates. They concluded that there is no benefit of this combined regimen in fresh IVF cycles and possible harm in frozen embryo cycles ^28^. The most contemporary Cochrane review of glucocorticoid steroid use specifically for peri-implantation support also found no evidence to support this practice ^9^. Among 16 clinical trials, this review found that steroids did not improve the rates of livebirth, clinical pregnancy nor miscarriage. In light of the lack of evidence of benefit, and the findings of our study that suggest harm, advocating for the ongoing use of periconceptual steroid use seems imprudent.

Previous studies have attempted to clarify the risk of congenital malformation associated with periconceptual steroid use. Carmichael et al reported in the late 1990s on the risk of specific congenital abnormalities following maternal corticosteroid exposure. They found that periconceptual steroid use was associated with a significant increase in the risk of cleft lip and palate abnormalities ^29^. In a meta-analysis of comparable studies, Park-Wyllie et al found that there was a marginally increased risk of major malformations after first-trimester exposure to corticosteroids, and a significantly increased risk of cleft abnormalities^18^. Other studies have not found an increase in cleft abnormalities but have reported significant increases in the risk of urogenital abnormalities^19^.

Past studies have been limited however by their small size (ie. 180-330 participants) ^18,30^, study design ^31–33^ and inability to identify and exclude patients with known indications for steroid use. One large population-wide cohort study in Denmark reported an association between topical corticosteroid use in early pregnancy and orofacial cleft abnormalities, but not steroids more broadly ^34^. Patients with an indication for steroid use were not excluded.

The proponents of periconceptual steroid use suggest their immune modulating effect suppress immune system abnormalities and improves endometrial receptivity – a critical aspect of successful pregnancy. However, this does not recognise that the immune system is critical to implantation and ongoing pregnancy, and does not need to be suppressed ^35^. Furthermore, it is plausible that the passage of prednisolone into the placental circulation, albeit small (~10%)^36^, could negatively impact the developing fetus, especially given the discovery of adverse outcomes in animal models^37^.

The strength of our study lies in the use of statewide linked data, which ensured an unbiased expansive cohort size and an ability to identify critical patient factors. Our study was designed *a priori* with input from a multidisciplinary team with expertise in fertility medicine, maternal-fetal medicine, pregnancy loss and epidemiology. The resulting study design and analysis is robust, with appropriate accounting for missing data and consideration of potential confounders.

Linkage of perinatal, prescribing and hospital admission data allowed us to ascertain pre-existing medical conditions warranting steroid use (and exclude these women), determine actual prescription and pharmacy dispensing of steroid medication and characterise specific outcomes. Our study design and scope thus ensured we were able to exclude patients with known indications for steroid use, and also identify specific congenital abnormalities, overcoming many of the limitations of past studies. Big data offers an opportunity in such circumstances, where clinical trials are limited by size, follow-up and legitimate ethical concerns about safety.

There are limitations, largely arising from the use of routinely collected observational data. Data on mode of conception are typically poorly recorded and this is likely to explain the low capture of ART-conception among the steroid-exposed cohort (4.8%). Despite this, our cohort numbers were large enough to allow sub-analysis of ART-only pregnancies to give an indication of the specific risk among this cohort. Missing data are inherent to observational cohorts. However, these data were minimal and where appropriate, imputed using the robust method of bootstrapping, an approach which offer benefits beyond an imputation-only approach^38^. Finally, our finding in our subgroup analyse of no increased risk of congenital abnormality following dexamethasone use should be viewed cautiously given the small sample size.

## Conclusion

Using population-wide, linked data our study has demonstrated that periconceptual steroid use is associated with an increased risk of major congenital malformations. In the absence of clear benefits, and now evidence of harm, periconceptual steroid use without a medical indication should be avoided for all patients, including those undergoing fertility treatment.

## Data Availability

Linked, de-identified administrative data are held in a secure online environment by the approved research team. Access to this data may be permitted following appropriate ethical approval and approval from all relevant data custodians.

## DECLARATIONS

### Availability of data and materials

The data that support the findings of this study are available from the corresponding author but restrictions apply to the availability of these data, which were used under license for the current study, and so are not publicly available. Data are however available from the authors upon reasonable request and with permission of the individual data custodians; The Consultative Counsel of Perinatal Mortality and Morbidity and the Australian Institute of Health and Welfare.

### Authors contributions

A.L and A.K had full access to all the data in the study and take responsibility for the integrity of the data and the accuracy of the data analysis. All authors qualify for authorship by contributing substantially to this article. A.L, A.K, R.H, S.W, S.T, B.V and C.S conceived the study. A.L and J.A completed the ethic applications, coordinated with the data custodians and linkage agency and performed the literature review. A.K performed the data cleaning and analysis. A.L wrote the manuscript and is the guarantor for this publication. R.J.H assisted in planning and performing the statistical analysis and contributed substantially to the STATA coding required for the analysis and performed sensitivity analysis. R.Ha, J.A, H.G, S.W, E.M and S.T provided intellectual support and guidance and assisted with preparation and writing of the manuscript. B.V, C.S and M.G provided expert guidance in the field of IVF. All authors have contributed to critical discussion, agreed upon the prespecified statistical analysis plan and reviewed and approved the final version of the article for publication.

## Acknowledgements

We are grateful to CCOPMM for providing access to the data used for this project and for the assistance of the staff at Safer Care Victoria. The conclusions, findings, opinions and views or recommendations expressed in this paper are strictly those of the author(s). They do not necessarily reflect those of CCOPMM.

